# Approach to some data preparation problems in systematic review and meta-analysis

**DOI:** 10.64898/2026.10.08.26365048

**Authors:** Richard J. Stevens, Francesca Little

## Abstract

In systematic review, the measures reported by the primary studies may not always be the measures sought by the systematic review investigators. Motivated by an systematic review of interventions to reduce meat consumption, we discuss here methods to infer relative risks and confidence intervals from randomized controlled trials that reported differences, possibly in the presence of repeated measures or multiple interventions.

## 1 Background

In systematic review and meta-analysis, between the systematic search for papers and a quantitative meta-analysis is data extraction. In some cases this is, literally, the extraction from a published paper of the numbers we wish to analyse. In other cases, some mathematical deduction is required to deduce the numbers needed for the analysis from the numbers published in the original report. The Cochrane Handbook describes common problems in data extraction, and methods to address them [1]. Our colleagues in the Centre for Evidence Based Medicine, University of Oxford, have also published a series of mathematical techniques that may be helpful in various situations [2][3].

We were recently asked by colleagues to assist with data extraction for a systematic review of interventions to reduce meat consumption. In the main analysis, the outcome of interest is relative risk, where “risk” is the probability that an individual chooses a meal containing meat. At the onset of our involvement, our colleagues had extracted the relative risk (or risk ratio, RR) for meat consumption from all included studies of two parallel groups in which each participant made a single choice. There were other studies that met the inclusion criteria but for which RR could not be calculated in this way. In this technical report, we describe the approach that we used, as statistical advisors to the systematic review authorship, to estimate RR and 95% confidence interval (CI) in such studies.

## 2 Approach to repeated measures binary outcomes

Suppose *n* participants in a trial are each observed on *k* occasions making a binary choice; let *Y*_*ij*_, *i* = 1, …, *n, j* = 1, …, *k* denote the observed choices, where *Y*_*ij*_ ∈ {0, 1}. If the participants are randomly allocated to two treatment groups, for investigators with access to patient-level data the RR and CI could be calculated by Binomial regression.

### Case 1

Such a trial may report the RR and CI from appropriate Binomial regression. In this case we extract that reported RR and CI for use in the meta-analysis. No further calculation is required.

### Case 2

Such a trial may report the mean and standard deviation (SD) number of successful trials. That is, having calculated 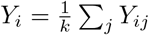 for each *i*, they may report the mean and SD of *Y*_*i*_ in each of two treatment groups.

In this case, we estimate RR by dividing the mean in the treatment group by the mean in the control group. Our approach to estimating a suitable CI for the RR is as follows: we seek a standard error (SE) for the logarithm of the RR (logRR) with the property that a Wald test of the hypothesis that logRR = 0 has a *p*-value consistent with the reported data. We operationalise this as follows.

Firstly, if a valid *p*-value, *p*, for difference between treatment and control groups has been published, we use this. We calculate the *z*-statistic corresponding to this *p*-value by solving

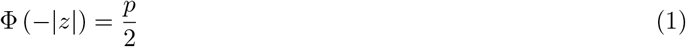

so that if *p* = 0.05, for example, then *z* = 1.96. This can be implemented in the *R* software[7] by

~~~
z <-abs(qnorm(p/2))
~~~

Then

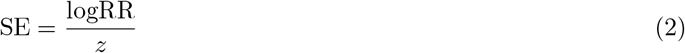

The standard error is then used to calculate a 95% confidence interval for logRR and hence for RR. r Secondly, if no valid *p*-value has been published, we use the reported mean, SD and sample size in each treatment group to conduct a *t*-test. This can be implemented in *R* by using the function tsum.test from the BSDA package [8]. For example, in the case of equal sized randomisation groups:

~~~
t<-tsum.test(mean.x=mean1, s.x=se1, mean.y=mean2, s.y=se2, n.x=n/2, n.y=n/2)$statistic
~~~

We then substitute *t* for *z* in equation (2) above.

## 3 Example

Stel [4] asked 561 participants to make 10 meal choices each. They reported the outcome as continuous, that is, reporting the mean and standard deviation (SD) times that participants chose a meal containing meat. Participants in the intervention group chose a mean of 8.22 (SD 5.36) times, and participants in the control group a mean of 9.13 (SD 5.31) times. We therefore estimate the RR for choosing meat in intervention vs. control to be 8.22*/*9.13 = 0.9003. To allocate a confidence interval, we note that a *t*-test on the continuous data yields a *t*-statistic of −2.02. For a log RR of log(0.9003) = −1.050, a standard error of 0.05198 is associated with a Wald statistic of −1.050*/*0.05198 = −2.02. We therefore use this standard error to obtain a confidence interval of (−0.207, −0.00312) for the log RR, and a 95% confidence interval of (0.813, 0.997) for the RR.

### Performance in simulated data

We simulated 16, 000 datasets of 560 participants, making 10 choices each, as in Stel’s study. The simulated participants in each dataset were randomly allocated to two equal size groups with probability 0.913 or 0.822 of choosing meat. Nominal 95% confidence intervals calculated by our method contained the true relative risk in 94.3% *±* 0.2% of simulations, where *±* denotes estimated standard deviation of Monte-Carlo error.

## 4 Approach to trials with multiple intervention groups

Suppose a trial randomises participants or centres to *k* intervention groups and 1 control group, where *k >* 1. For simplicity, we consider here the case of approximately equal sized intervention groups; this was sufficient for the review that motivated the current report. The Cochrane Handbook [5] discusses several options. The first is to combine all intervention groups into one, yielding a single comparison for inclusion in the meta-analysis. The second is to retain the individual comparisons, but with the control group “divided out approximately evenly” among the comparisons. A third is to account for the correlation between comparisons mathematically. The final suggestion is network meta-analysis: we rule this out as beyond the scope of the current review.

Whether to combine intervention groups, or retain the individual comparisons, is a matter for the protocol of the systematic review, and beyond our scope here. In circumstances when the individual comparisons are to be retained, we take the following approach. Where the study design and the reported data permit, we take the “divided” control group approach. For example, in studies with a single binary observation per participant we divide the number of events and number of participants in the control group by *k*, and then calculate the relative risk and 95% confidence interval.

In other cases (for example, the repeated measures context described in Section 2) this simplistic approach does not apply because the relative risk is not calculated simply from the number of events and number of participants. In that case, our approach is to inflate the standard error of each comparison to reflect the correlation. Borenstein et al. [6] note that for two equally-sized intervention groups and one control group, the expected correlation between the two comparisons is ^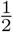^. More generally, for *k* equally-sized intervention groups compared to one control group, the expected correlation between any two comparisons against the control group is 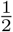. If we enter the *k* comparisons into a fixed effect meta-analysis, the comparisons as a whole will be over-weighted in the meta-analysis by a factor of 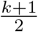 (see Appendix 1).

When retaining all *k* comparisons therefore we inflate standard errors by a factor of 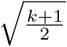. This has the property that in a fixed effect meta-analysis, the overall weight allocated to the study is approximately the same as if the study had been reported as a single comparison of (any intervention) vs. (control).

For a continuous outcome, if the control group is approximately the same size as the intervention groups and has approximately the same standard deviation, then this inflation factor is approximately the same as the “divided” control group approach. This also holds for a binary outcome if the effect is close to null (see Appendix 2).

## 5 Discussion

The method in Section 2 is suggested as a common-sense approach to non-linear transformation of an effect measure. By preserving the statistical “significance” of a finding, we hope to maintain an appropriate relative weight for the study in the resulting meta-analysis. This work being unresourced, we have not been able to comprehensively investigate the literature for precedent - it may be that this method has been previously used and/or previously investigated.

The simulation in Section 3 is limited in scope. Further investigations should consider a range of baseline probabilities, relative risks and sample sizes. Coverage of confidence intervals is not necessarily the best metric: an alternative would be to estimate RR and confidence interval within each dataset by a repeated measures Binomial model, and compare the resulting Wald statistic directly to the *t*-statistic from the continuous analysis. However, there are pragmatic difficulties since algorithms for repeated measures Binomial models are not guaranteed to converge on every dataset.

Since the method in Section 4 depends on approximating assumptions, particularly for binary outcomes, we suggest that the long-established approaches of combining the intervention groups or using a “divided control group” be preferred where possible.

Our approach throughout gives emphasis to the weighting that arises in fixed effect meta-analysis. There are other approaches to meta-analysis that use the same weighting scheme, including the Henmi and Copas approach to random effects meta-analysis [9], and the Inverse Variance Heterogeneity method, which may be interpreted either as a random effects method or as a fixed effect method with adjustment for overdispersion [10]. However, most methods for random effects meta-analysis incorporate the estimated between-study variance into the weighting scheme. The logic of fixed effect meta-analysis will apply approximately at best, when heterogeneity is low.

## Data Availability

The data used in the manuscript are in the public domain.

## Acknowledgments

GPT-5.6 Sol, accessed through the University of Oxford ChatGPT Edu installation, was used to check the manuscript for mathematical or typographical errors. Any remaining mistakes are our own.

## Appendix 1

Suppose *k* effect parameters *β*_1_, …, *β*_*k*_ represent the comparisons of *k* intervention arms to a common control group. Then

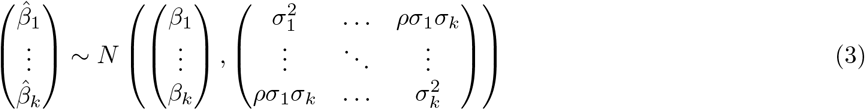

With equal-sized randomization groups, 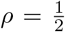. Suppose the *k* values of 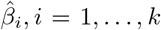 are entered into a fixed effect meta-analysis, weighted by the inverse variances, as if *ρ* = 0. We obtain a weighted average 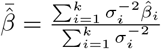.

The fixed effect meta-analysis formula for the variance of the weighted average is

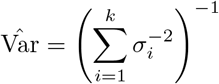

Under the approximating assumption that all variances are equal, *σ*_*i*_ = *σ∀i*, this reduces to

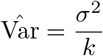

However, it follows from (3) that, under the equal variances assumption, the actual variance of 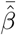 is

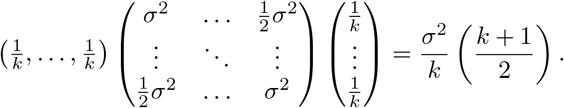

## Appendix 2

Let *X*_*i*_, *i* = 1, …, *n*_*x*_ be the values of a continuous outcome in *n*_*x*_ individuals randomised to treatment, and let *Y*_*i*_, *i* = 1, …, *n*_*y*_ be the values of a continuous outcome in *n*_*y*_ individuals randomised to control. The variance of the difference in means is

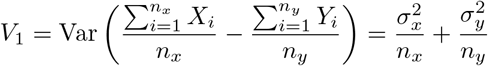

where 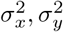 are the variances in the two groups. Now suppose investigators artificially divide the control group into *k* equal sized groups, as described above. Then the variance of the difference in means between the intervention group and the “divided” control group is

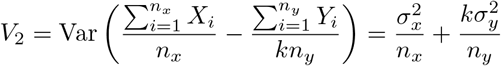

Suppose 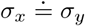 and 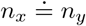. Then 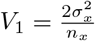 and

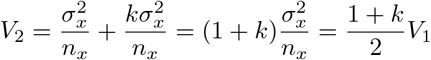

Alternatively, suppose *a* of *a* + *b* observations in a treatment group, and *c* of *c* + *d* observations in a control group, have the outcome of interest. The usual estimate of the variance of the log odds ratio is

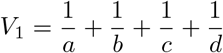

If investigators artificially divide the control group into *k* equal sized groups, then the resulting variance estimate is

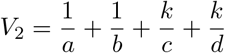

If the two groups are of equal size, so that (*a* + *b*) = (*c* + *d*), and if the odds ratio is approximately 1 so that 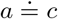 and 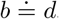,

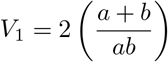

and

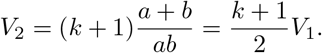

## Notes

### Competing Interest Statement

The authors have declared no competing interest.

